# Rapid assessment of the impact of “lockdown” on the COVID-19 epidemic in Portugal

**DOI:** 10.1101/2020.05.26.20098244

**Authors:** Vasco Ricoca Peixoto, André Vieira, Pedro Aguiar, Carlos Carvalho, Daniel Thomas, Alexandre Abrantes, Public Health Research Center, National School of Public Health, Portugal

## Abstract

**Background:** Portugal took early action to control the COVID19 epidemic, imposing a lockdown on March 16 when it recorded only 62 cases of COVID-19 per million inhabitants and no reported deaths. The Portuguese people complied quickly, reducing their overall mobility by 80%. We estimate the impact of the lockdown in Portugal in terms of reducing burden on the health service.

**Methods:** We forecasted epidemic curves for: Cases, hospital inpatients (overall and in ICU), and deaths without lockdown, assuming that the impact of containment measures would start 14 days after lockdown was implemented. We used exponential smoothing models for deaths, intensive care (ICU) and hospitalizations and an ARIMA model for number of cases. Models were selected considering fitness to the observed data to the 31st of March 2020. We then compared observed(with intervention) and forecasted curves (without intervention).

**Results:** Between April 1 and April 15, there were 146 fewer deaths(−25%), 5568 fewer cases (−23%) and, as of April 15, there were 519 fewer ICU inpatients(−69%) and 508 fewer overall hospital inpatients(−28%) than forecasted without lockdown. On April 15 the number of ICU inpatients could have reached 748, three times higher than the observed value (229) if the intervention had been delayed.

**Conclusion:** If the lockdown had not been implemented in mid-March, Portugal ICU capacity (528 ICU beds) would likely have been breached in the first half of April. The lockdown seems to have been effective in reducing transmission of SARS-Cov-2, serious Covid-19 illness and associated mortality, thereby decreasing demand on health services. Early action allowed time for the National Health Service to acquire protective equipment, to increase capacity to test and cope with the surge in hospital and ICU demand caused by the pandemic.

## Introduction

Since there is no vaccine or treatment for COVID-19, governments have used social and behavioural interventions to reduce spread of the virus in the community. Recent studies suggest that these public health measures have had an impact. Whilst individual measures, for example: contact tracing and isolation of cases and contacts, wearing masks, movement restrictions and other measures to reduce social contacts and physical proximity may have an impact^1^, it has been suggested that only through a combined set of measures can the spread of the virus be contained. ^2 3 4 5^

The European Centre for Disease Prevention and Control (ECDC), in a technical report “Strategies for Surveillance”^3^ recommends that the effectiveness of containment measures should be assessed at regular intervals by monitoring intensity, and the impact on the healthcare system. The report also stresses the importance of frequent, open and transparent communication with the public to explain these findings, in order for the population to accept and comply with the chosen mitigation measures over an extended period of time.

Portugal took early action to control the COVID-19 epidemic, imposing restrictions on economic activity and social life when there were only 62 cases of COVID-19 per million inhabitants and no COVID-19 deaths, a different epidemiological situation than that of Spain, Italy and the United Kingdom, when equivalent measures were taken later in the course of the epidemic^5^. International comparison of the ‘Stringency Index’, a summary score taken from 17 indicators of government responses compiled by the Oxford COVID-19 Government Response Tracker,^6^ indicates that Portugal implemented in mid-March stringent containment and mitigation measures, including the cancellation of public events, school closures, workplaces and restriction of national and international movement.

As the Stringency Index increased and lockdown was implemented, the Portuguese people complied with these confinement measures and quickly reduced their overall mobility, (Figure 1.) According to data published by Google^7 8^ and Apple^9 5^ the Portuguese people reduced significantly their daily mobility, including for retail and leisure (−83%), parks and alike (−80%) and transport (−79%)^8^. The population in Spain also adhered effectively to government containment and mitigation measures. In Italy and UK, on the other hand, the population seems to have been slower reduction in mobility as the stringency increased eventually reflecting different communication and risk perception.^5^

**Figure 1.**
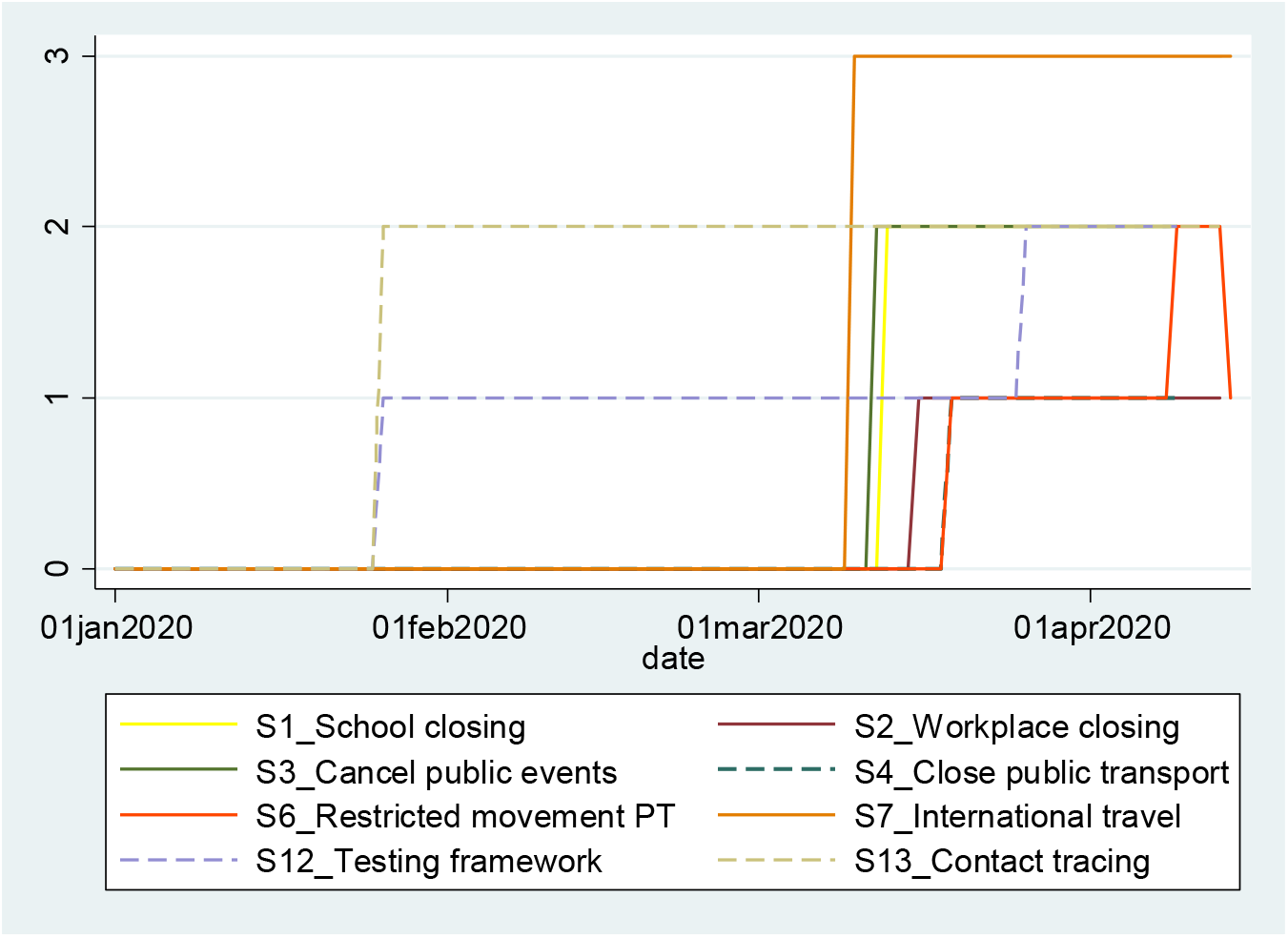
Variation in selected Indicators of the Oxford Government Response Stringency Index in Portugal, 2020

**Figura 1.**
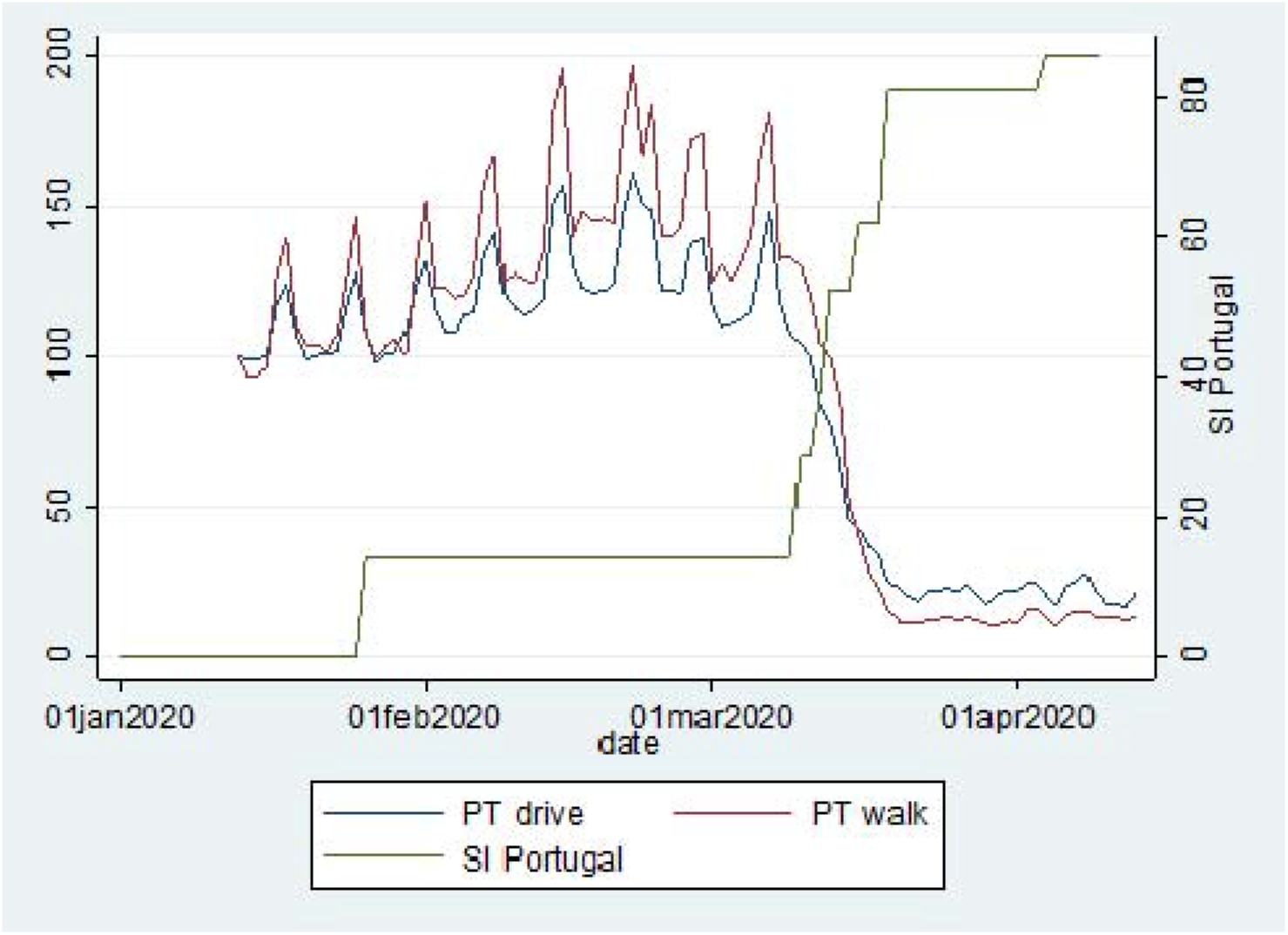
Temporal trend in the Oxford Contingency Index (green) and mobility by car(blue) and walking (red) as defined by Apple Mobility, Portugal: January 13 to April 15.

In March 2020,187 people died of COVID-19, or 2.3% of the 8 521 confirmed cases, a cumulative incidence of around 80 cases per 100 000 inhabitants and a mortality rate of 2.3%.

In line with one of the ECDC strategies for surveillance of COVID-19, and the WHO COVID-19 strategy recommendations on research and sharing knowledge^10^ this paper estimates the early direct health impact of the lockdown in Portugal, that is: on the number of COVID-19 cases, deaths and clinically severe cases (using number of hospital or intensive care unit beds occupied as proxy indicator of serious illness).

## Methods

Data on the daily number of cases, deaths, and prevalent number of cases in patients in hospital and in intensive care (ICU) were collected from official, publicly available^11^ COVID-19 Situation Reports of the Directorate-General of Health of Portugal until April 15, 2020, also available through the ECDC (cases and deaths). For data on hospitalization and ICU attendance, occupied hospital beds (overall) and ICU beds, rather than new admissions, were used as indicators of prevalent hospitalized Covid-19 cases.

Firstly, we estimated the daily number of COVID-19 cases and associated deaths, hospital and ICU in April 2020 that would have occurred without containment measures. This was carried by forecasting values for April using exponential smoothing and ARIMA models selected based on fitness to the values recorded between March 1 and 31. Then, we compared the number of COVID-19 patients present in ICU and hospital each day to the number expected. For these outcomes we compared the values observed and expected on April 15 as a cumulative measure. We used SPSS expert modeler to consider different types of exponential smoothing and ARIMA models for specific time-series^12^, and find the best fitting models for the time series to March 31. Forecasts were obtained with exponential smoothing models applied to the time series of daily deaths, hospitalized in ICU, and total hospitalized patients to March 31. An ARIMA model was applied to the time-series of new cases, due to a better adjustment of the model. All models had good adjustments to the time-series until March 31, as demonstrated by the parameters meters presented in results. The analysis was performed in SPSS 26 using the approach described by B. Tabachnich for traditional models forecast^13^.

We considered a delayed effect of measures starting 14 days after the lockdown considering different pieces of evidence on the period from infection to onset of symptoms to the detection of cases, hospitalization (general ward or intensive care unit), and death. As such, since reduction in mobility and contacts between citizens was effective in mid-March 2020 we modelled observed data to March 31.(Figure 1.)

## Results

### Impact in daily deaths

In the analysed period, there were 442 deaths from COVID-19, 146 (−25%) fewer than the 588 that would be expected for that period if no containment and mitigation measures had been implemented. The exponential smoothing model for deaths (until March 31) had a good fit to observed data R2=0.91, smoothing parameter test p<0.001, quality adjustment Ljung Box P=0.75, FAC and FACP not significant (Figure 2.).

**Figure 2.**
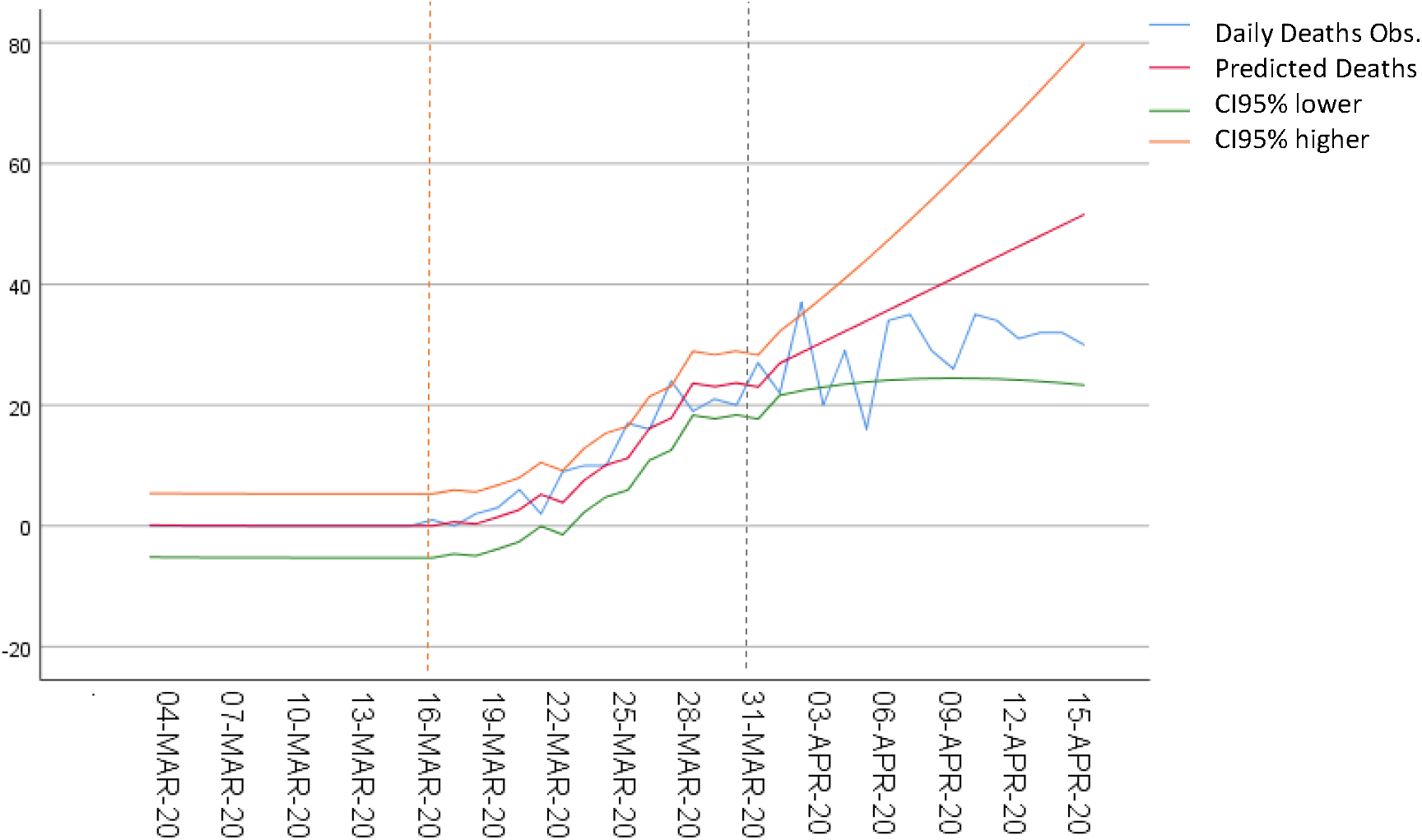
Observed and predicted n<u>^o^</u> of daily deaths by COVID-19, with 95% confidence levels (orange dashed line- date of lockdown; dashed grey line- beginning of forecast)

### Impact on ICU inpatients

Using publicly available data on prevalent hospitalized cases(overall) and ICU cases in each day, the forecast showed that as of April 15, 748 patients would be occupying ICU beds. We observed 519 fewer patients in ICU than the predicted value by that date (−69%). ICU bed occupation fell short of the lower bound of the 95% confidence interval generated by the model throughout the period.

Throughout the period form 1 to 15 April, there was a daily average of 237 COVID-19 occupied ICU beds, 269 fewer than the 506 daily average expected in the same period (−53%), without containment and mitigation measures. For this analysis, we used an exponential smoothing model of number of patients in ICU (March 31), R2=0.98, p<0.001 smoothing parameter test, Ljung Box adjustment quality P=0.96, FAC and FACP not significant (Figure 3.).

**Figure 3.**
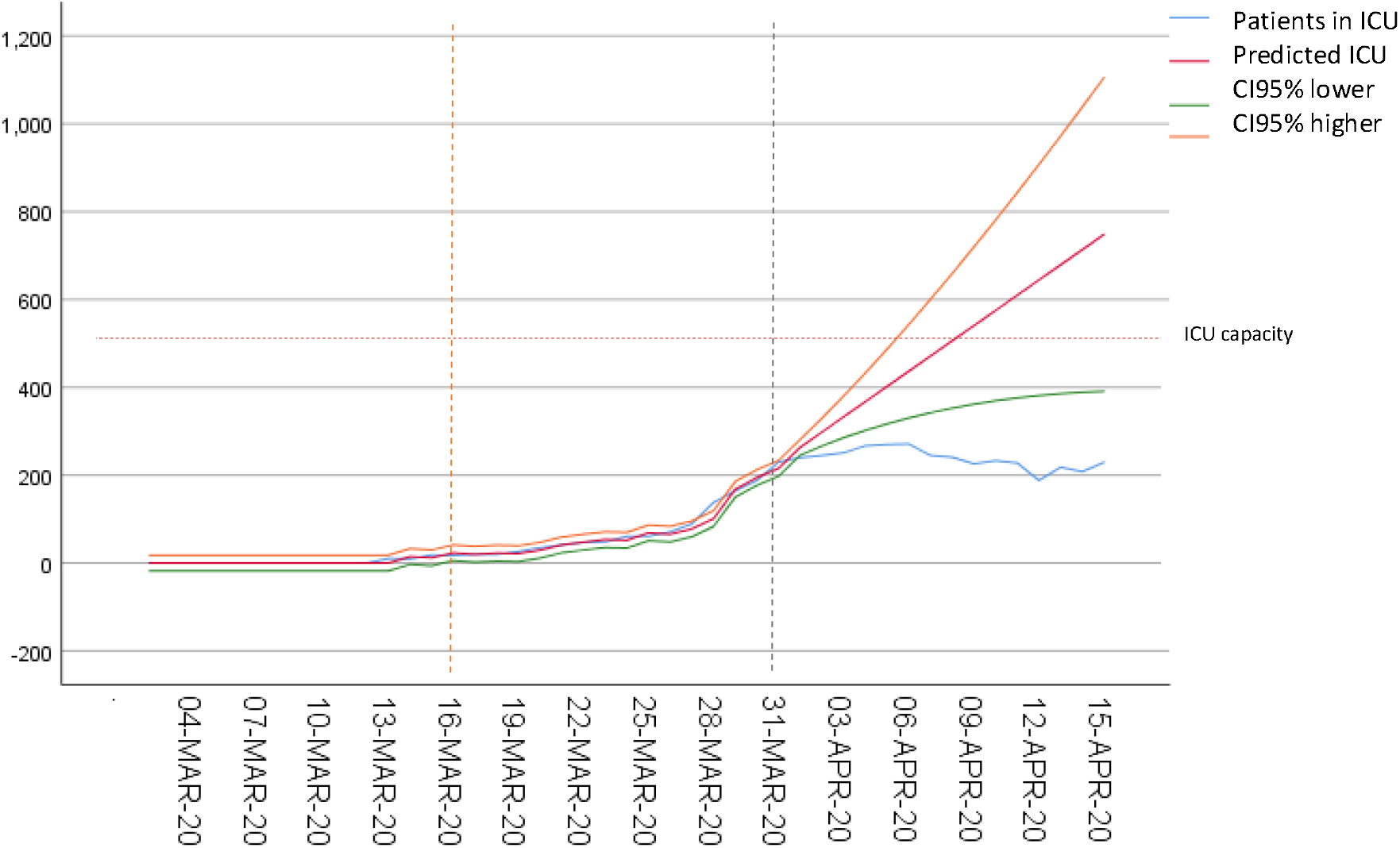
Observed and predicted n<u>^o^</u> of daily ICU inpatients with COVID-19, with 95% confidence levels (orange dashed line- date of lockdown; dashed grey line- beginning of forecast; Red dashed line -ICU beds capacity:528)

### Impact on overall hospital beds occupation

We used publicly available data on occupied hospital beds(overall) each day to model the forecast. As at April 15, we predicted 1810 overall hospital beds occupied. We observed 508 fewer than the predicted value for that date (−28%).

Between 1 and 15 April, there was a daily average of 1158 <u>hospital beds occupied</u> by COVID-19 patients, 142 fewer than the 1300 occupied beds expected (−11%) if no containment and mitigation measures had been put in place. For this analysis we used the exponential smoothing model of hospitalized patient numbers (to March 31), R2=0.94, smoothing parameter test p<0.001, Ljung Box P=0.84 adjustment quality, FAC and FACP not significant(Figure 4.).

**Figure 4.**
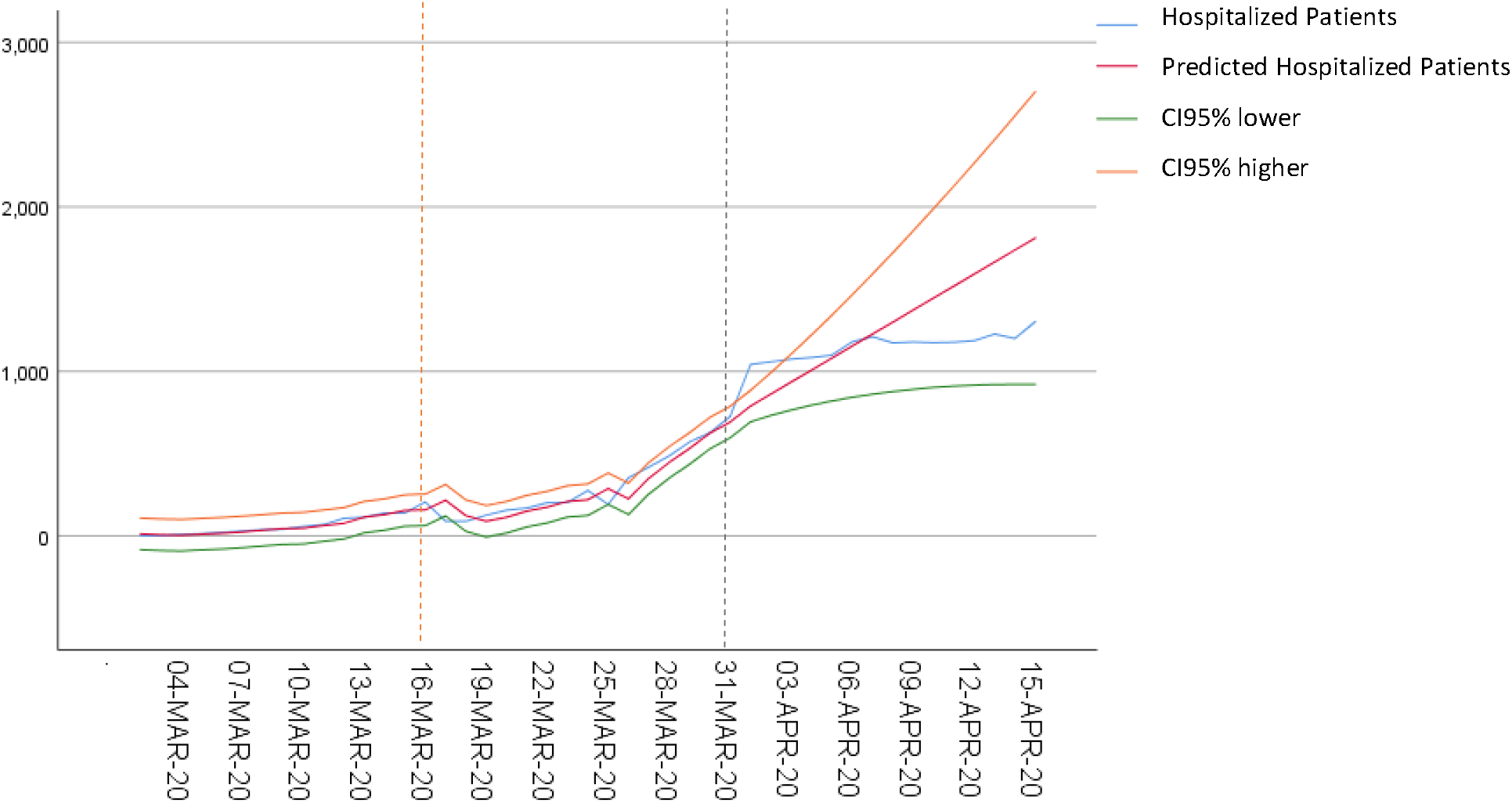
Observed and predicted n<u>^o^</u> of daily hospital inpatients (All) with COVID-19, with 95% confidence levels (orange dashed line- date of lockdown; dashed grey line- beginning of forecast)

### Impact in daily new cases

Between 1 and 15 April, there were 5568 fewer cases than the 24405 cases forecasted (−23%). This indicator remained under the lower bound of the 95% confidence interval generated by the model after April 9.The forecast used an ARIMA model (2,1,0) adjusted until March 31 for the number of new daily cases 1, R2=0.86, test parameters of the model p<0.05, adjustment quality Ljung Box P=0.95, FAC and FACP not significant (Figure 5.)

**Figure 5.**
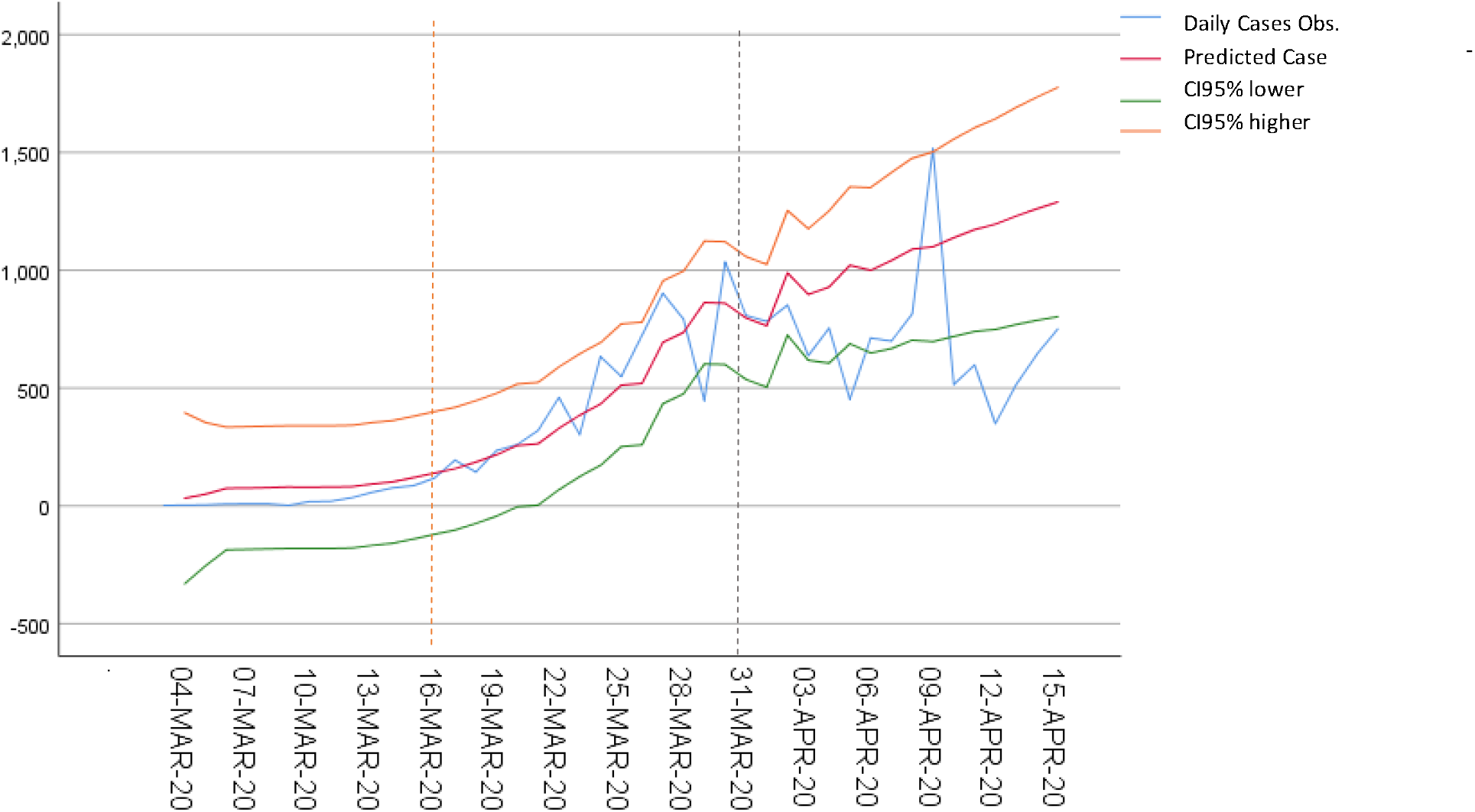
ARIMA model. Daily n<u>^o^</u> of observed and predicted n<u>^o^</u> of cases of COVID-19 and 95% confidence intervals. Orange dashed line - date of lockdown; dashed grey line- beginning of forecast.

**Table 1.** Predicted and observed values and absolute and relative differences for different COVID-19 indicators from April 1^st^ to 15^th^

|  |  | Predicted | Observed | Dif | Dif% |
| --- | --- | --- | --- | --- | --- |
| **Deaths** | Average number of daily Deaths | 39.23 | 29.47 | -9,76 | -0.25% |
|  | Total Deaths | 588 | 442 | -146 | -0.25% |
| **Patients in ICU** | Average number of occupied beds | 237.33 | 505.7517 | -268 | -53% |
|  | Total ICU inpatients on April 15 | 229 | 748 | -519 | -69% |
| **All Hospitalized Patients** | Average number of inpatients | 1157.93 | 1299.689 | -142 | -11% |
|  | Total inpatients on April 15 | 1302 | 1810 | -508 | -28% |
| **Cases** | Average number of daily new cases | 567 | 428 | -139 | -25% |
|  | Total cases on April 15 | 24405 | 18837 | -5568 | -23% |

## Discussion

The findings of this study suggest that early Government action in implementing a strict containment and mitigation policy and a high level of compliance of the Portuguese population were effective in reducing mortality and severe morbidity of COVID-19. Between April 1 and April 15, there were 25% fewer deaths, 23% fewer cases and, as of April 15, there were 69% fewer ICU inpatients and 28% fewer overall hospital inpatients than expected. On April 15 the number of ICU inpatients could have been greater than 740, more than 3 times higher than the observed value if the intervention was delayed beyond the end of March assuming a 14 day lag in impact.

These time-series forecasting methods allow for an early retrospective estimate of the impact of measures that may be repeated whenever containment measures are changed and give an intuitive way to visualize the impact of interventions. They are adequate for short-term forecasting making quantitative projections for policy makers^14^ that are of relevance to public communication when justifying control measures^3^. However, they do not consider changes in parameters governing transmission, disease outcomes, and immunity to predict outcomes in the long-term as is done by mechanistic modelling^14^.

Despite being useful, there are some limitations in the quality of surveillance data we used in these models. Early in the epidemic there was likely to be an under-ascertainment of cases leading to our forecast being more conservative. On the other hand there may have been some delays in reporting, for example, resulting in a peak in reporting of cases on April 9 which is otherwise unexplained.

Data on Covid-19 deaths, and occupied Hospital (overall) and ICU beds are of reasonable quality. In Portugal, COVID-19 deaths are reported by clinicians using an online national platform and these data are available in real time. Deaths in patients who were suspected cases of COVID-19 where a lab result was not available are tested post-mortem^15^ (suspect case definition stopped including epidemic link with a confirmed case on March 26). Prevalent ICU and overall hospitalized cases in each day are reported from each hospital to the Regional level and to DGS that collates and communicates the data and as such we assume a high level reporting quality. However, data on new cases are likely to be biased, reflecting testing strategies and tests availability. There was likely to be a high level of under ascertainment in the early phase of the epidemic. As the testing strategy changed in Portugal from March 26 and testing became more widespread (everyone with cough or fever tested).

Our model made a number of assumptions based on data presented in the published literature, which varies ^16 17 18^. One study found the median incubation period of COVID-19 is 7 days (IQR:4-11)^16^, another that the median time from first symptom to dyspnoea being 5 days, to hospital admission was 7 days, and to ARDS was 8 days ^17^; an interrupted time-series study suggest that the onset of reduction effects after COVID-19 lockdown in Hubei and Guandong on incidence and mortality were observed after a period ranging from 7 to 17 days and 10 days, respectively^19^. Considering this and for and easier reading and interpretation we assumed in the analysis that the impact would begin to be observed for all the outcomes from April 1,14 days after the lockdown.

Further supporting the decision to use 14 days as a cut-off, according to the National Association of Public Health Physicians (ANMSP), R (t) in Portugal (considering the previous 7 days) has rapidly decreased from 3.64 in March 18, to 2.2 in March 24 to 1.64 in March 30 being almost always less than 1 since April 6.^20^ Our estimates are conservative. This shortterm forecasting method assumed a fixed cut-off date on March 31,14 days after lockdown, to start forecasting the number of deaths, hospital and ICU inpatients and cases without intervention. The impact of the lockdown measures however must have started earlier and gradually, rather than on specific moment in time.^19 21^ The gradual reduction in R(t) in Portugal corroborates this. However, since R(t) and mobility reduction happened quickly in Portugal, by the middle of March, the effect would still not be too spread over time. Our time series models incorporate a flattening of the new cases and death curves which was already happening in the last days of March 2020. This influences the forecasts making them more conservative for these outcomes

We cannot isolate the effect of specific measures on different outcomes. ICU cases and deaths are more strongly influenced by the number of cases in elderly population, since they have a higher risk^22^ and may have a larger impact if more cases are prevented in this population.

Different methods have been used internationally for estimating impact of COVID-19 containment measures through Susceptible Infected and Recovered models and others ^23 24 25 26 27^ including retrospectively through interrupted time-series ^19 21^. The latter consistently found an impact of lockdown policies, with variable lags from lockdown to maximal impact.

We believe this forecast to be adequate even if conservative as some behavioral change would occur even without severe lockdown measures.

The timing of the implementation of strict social distancing varied in different European countries. In Portugal implementation was relatively early, following lessons learned from earlier experience in Italy and Spain. Early interventions may have a been particularly effective in early phases of the pandemic were a large proportion of mild cases may have accumulated undetected^28 29^ and under-ascertainment estimates vary widely in different countries ^30 31^.

The population risk perception may have been influenced early by media reports on neighboring countries. Indeed a Social Opinion periodic survey of a non probabilistic sample with more than 150.000 respondents found that risk perceptions were high from the week starting in March 21 (20,6% high risk; 44,9% moderate risk of acquiring COVID-19) and remained high until the end of the first weak of April, with slight reduction afterwards. ^32^

This type of modelling work, alongside other methods can be reproduced to find early evidence for impact of changing containment strategies when lifting stringent social distancing policies although decisions of modeled baseline period and initiating forecast can be further discussed in different setting considering how quickly policy decisions have effective impact in behavior(mobility measures from Google and Apple should be included in surveillance), new infections and severe outcomes of infection.

## Conclusions

In Portugal, early and quick containment measures and high level of compliance of the population were associated with a relevant reduction in the number of serious cases and deaths by COVID-19. Results were apparent less than two weeks after lockdown. This may have bought time for preparedness and response to allow for the implementation of other measures, including acquisition of protective equipment and increasing health-care capacity.

The capacity of the National Health Service to care for serious COVID-19 cases, (528 intensive care unit beds at the start of the epidemic, would likely have been breached if containment measures had been delayed to the end of March. In may, ICU beds capacity had been increased to 713 according to the Health Ministry ^33^. On other note, Regional spread has been heterogeneous in Portugal^11^ making it more likely that ICU capacity would have been breached earlier in Regions most affected.

As for relaxation in lockdown measures Portugal was in the beginning of May in a good position to start easing social distancing, relaunching economic and social life. However risk communication strategies must be in place to guarantee compliance with other prevention measures of social distancing, mask wearing, respiratory and hand hygiene since the Portuguese population felt a smaller first wave epidemic. During this period, it is necessary to maintain a high level of epidemiological surveillance, to detect early and precisely any new surge of the epidemic, b) focus on keeping the number of serious cases and deaths down, focusing on the high risk population (people aged 70+, and those with debilitating illness, namely those in long-term care institutions), c) protect health care workers and other high risk professions, c) keep the transmission rate under control, as recommended in ECDC Risk Assessment of April 9^34^ and the European Comission^35^.

## Data Availability

All data is publicly available from official Portuguese Directorate-General of Health

https://covid19.min-saude.pt/relatorio-de-situacao/

## Key points

- Early containment measures and lockdown in Portugal were effective in controlling the epidemic and may have allowed ICU capacity to cope with demand that could have been surpassed by April 15, if lockdown was delayed;
- This may have bought time for preparedness and response to allow for the implementation of other measures, including acquisition of protective equipment and increasing health-care capacity.

## References

1. Koo JR, Cook AR, Park M, et al. Interventions to mitigate early spread of SARS-CoV-2 in Singapore: a modelling study. Lancet Infect Dis. March 2020. doi:10.1016/s1473-3099(20)30162-6

2. The effect of non-pharmaceutical interventions on COVID-19 cases, deaths and demand for hospital services in the UK: a modelling study | CMMID Repository. https://cmmid.github.io/topics/covidl9/control-measures/uk-scenario-modelling.html. Accessed April 6, 2020.

3. European Centre for Disease Prevention and Control. Strategies for the surveillance of COVID-19. Stockholm: ECDC; 2020. https://www.ecdc.europa.eu/en/publications-data/strategies-surveillance-covid-19. Accessed April 16, 2020.

4. Kraemer MUG, Yang C-H, Gutierrez B, et al. The effect of human mobility and control measures on the COVID-19 epidemic in China. Science (80-). 2020;4218(March):eabb4218. doi:10.1126/science.abb4218

5. Ricoca Peixoto V., Vieira A., Aguiar P., Sousa P., Abrantes A. “Timing”, Adesão e Impacto Das Medidas de Contenção Da COVID-19 Em Portugal / COVID-19. https://covid360.unl.pt/?p=1595. Accessed May 9, 2020.

6. Oxford COVID-19 Government Response Tracker | Blavatnik School of Government, https://www.bsg.ox.ac.uk/research/research-projects/oxford-covid-19-government-response-tracker. Accessed April 7, 2020.

7. Google. COVID-19 Community Mobility Reports. https://www.google.com/covidl9/mobility/. Accessed April 4, 2020.

8. Barómetro Covid-19. Mobilidade dos Portugueses à lupa da Google e do Barómetro Covid-19. https://barometro-covid-19.ensp.unl.pt/mobilidade-dos-portugueses-a-lupa-da-google-e-do-barometro-covid-19/. Accessed April 24, 2020.

9. Apple. COVID-19 - Mobility Trends Reports. https://www.apple.com/covid19/mobility. Accessed April 18, 2020.

10. WHO. COVID-19 strategy update -14 April 2020. https://www.who.int/publications-detail/covid-19-strategy-update-14-april-2020. Accessed May 15, 2020.

11. Direção Geral da Saúde. Relatónos de Situação - COVID-19. https://covid-19.min-saude.pt/relatorio-de-situacao/. Accessed May 15, 2020.

12. IBM Knowledge Center. IBM SPSS Time Series Modeler. https://www.ibm.eom/support/knowledgecenter/SSLVMB_24.0.0/spss/trends/idh_idd_tab_vars.html. Accessed May 12, 2020.

13. Tabachnick BG, Fidell LS. Using Multivariate Statistics (6th Ed.).; 2012. doi: 10.1037/022267

14. Holmdahl I, Buckee C. Wrong but Useful — What Covid-19 Epidemiologic Models Can and Cannot Tell Us. N Engl J Med. May 2020:NEJMp2016822. doi:10.1056/NEJMp2016822

15. Direção-Geral da Saúde. Norma n° 002/2020 de 16/03/2020 atualizada a 19/03/2020 Infeção por SARS-CoV-2 (COVID-19)—Cuidados post mortem, autópsia e casas mortuárias Normas - COVID-19. https://covid19.min-saude.pt/normas/. Accessed May 18, 2020.

16. Wang P, Lu J, Jin Y, Zhu M, Wang L, Chen S. Statistical and network analysis of 1212 COVID-19 patients in Henan, China. Int J Infect Dis. April 2020. doi: 10.1016/j.ijid.2020.04.051

17. Wang D, Hu B, Hu C, et al. Clinical Characteristics of 138 Hospitalized Patients with 2019 Novel Coronavirus-Infected Pneumonia in Wuhan, China. JAMA - J Am Med Assoc. 2020. doi:10.1001/jama.2020.1585

18. Yang X, Yu Y, Xu J, et al. Clinical course and outcomes of critically ill patients with SARS-CoV-2 pneumonia in Wuhan, China: a single-centered, retrospective, observational study. Lancet Respir Med. 2020;8(5):475–481. doi:10.1016/S2213-2600(20)30079-5

19. Medeiros de Figueiredo A, Daponte Codina A, Moreira Marculino Figueiredo DC, Saez M & Cabrera León A. Impact of lockdown on COVID-19 incidence and mortality in China: an interrupted time series study. [Preprint]. Bull World Health Organ. E-pub: 6 April 2020. doi: http://dx.doi.org/10.2471/BLT.20.256701.

20. ANMSP. COVID-19 | Mapa Epidemiológico Portugal. https://www.anmsp.pt/covid19-mapa. Accessed April 24, 2020.

21. Siedner MJ, Harling G, Reynolds Z, Gilbert RF, Venkataramani A, Tsai AC. Social distancing to slow the U.S. COVID-19 epidemic: an interrupted time-series analysis. *medRxiv*. April 2020:2020.04.03.20052373. doi: 10.1101/2020.04.03.20052373

22. Verity R, Okell LC, Dorigatti I, et al. Estimates of the severity of coronavirus disease 2019: a model-based analysis. Lancet Infect Dis. 2020;3099(20):1–9. doi:10.1016/S1473-3099(20)30243-7

23. Leung K, Wu JT, Liu D, Leung GM. First-wave COVID-19 transmissibility and severity in China outside Hubei after control measures, and second-wave scenario planning: a modelling impact assessment. Lancet. 2020;395(10233): 1382–1393. doi:10.1016/S0140-6736(20)30746-7

24. Lai S, Ruktanonchai NW, Zhou L, et al. Effect of non-pharmaceutical interventions for containing the COVID-19 outbreak: an observational and modelling study. *medRxiv*. March 2020:2020.03.03.20029843. doi: 10.1101/2020.03.03.20029843

25. Kucharski AJ, Russell TW, Diamond C, et al. Early dynamics of transmission and control of COVID-19: a mathematical modelling study. Lancet Infect Dis. 2020;0(0). doi:10.1016/S1473-3099(20)30144-4

26. Hellewell J, Abbott S, Gimma A, et al. Feasibility of controlling COVID-19 outbreaks by isolation of cases and contacts. Lancet Glob Heal. 2020;0(0). doi:10.1016/S2214-109X(20)30074-7

27. WHO Collaborating Centre for Infectious Disease Modelling; MRC Centre for Global Infectious Disease Analysis; Abdul Latif Jameel Institute for Disease and Emergency Analytics; Imperial College London. Impact of non-pharmaceutical interventions (NPIs) to reduce COVID-19 mortality and healthcare demand. 2020. https://www.imperial.ac.uk/mrc-global-infectious-disease-analysis/news--wuhan-coronavirus/. Accessed March 17, 2020.

28. Ricoca Peixoto V, Nunes C, Abrantes A. Epidemic Surveillance of Covid-19: Considering Uncertainty and Under-Ascertainment. Port J Public Heal. April 2020:1–7. doi: 10.1159/000507587

29. Munster VJ, Koopmans M, van Doremalen N, van Riel D, de Wit E. A Novel Coronavirus Emerging in China — Key Questions for Impact Assessment. N Engl J Med. January 2020. doi:10.1056/nejmp2000929

30. Center for Global Infecitous Disease Analysis - Imperial College London. Short-term forecasts of COVID-19 deaths in multiple countries. https://mrc-ide.github.io/covid19-short-term-forecasts/index.html#analysis-of-trends-in-reporting. Accessed May 13, 2020.

31. Russel T, Hellewell J, Abbott S, et al. Using a delay-adjusted case fatality ratio to estimate under-reporting. C Repos. 2020:1–6. doi:10.5281/ZENODO.3635417

32. Barómetro Covid-19-Opinião Social — 1 mês de confinamento: o que mudou nas perceções dos portugueses? https://barometro-covid-19.ensp.unl.pt/1-mes-de-confinamento-o-que-mudou-nas-percecoes-dos-portugueses/. Accessed May 18, 2020.

33. Diário de Notícias. Portugal nunca utilizou a sua capacidade total em cuidados intensivos. https://www.dnoticias.pt/pais/portugal-nunca-utilizou-a-sua-capacidade-total-em-cuidados-intensivos-CF6241613. Accessed May 9, 2020.

34. Coronavirus disease 2019 (COVID-19) in the EU/EEA and the UK—ninth update, 23 April 2020. Stockholm: ECDC; 2020. https://www.ecdc.europa.eu/en/publications-data/rapid-risk-assessment-coronavirus-disease-2019-covid-19-pandemic-ninth-update. Accessed April 30, 2020.

35. European Commission. A European roadmap to lifting coronavirus containment measures. https://ec.europa.eu/info/live-work-travel-eu/health/coronavirus-response/european-roadmap-lifting-coronavirus-containment-measures_en. Accessed April 20, 2020.

